# Clinical trial and real-world data: A comparative study in patients with diabetic kidney disease

**DOI:** 10.1101/2023.06.16.23291441

**Authors:** Samu Kurki, Viivi Halla-aho, Manuel Haussmann, Harri Lähdesmäki, Jussi Leinonen, Miika Koskinen

## Abstract

**Objective:** A growing body of research is focusing on clinical real-world data (RWD) to supplement or replace randomized controlled trials (RCTs). However, due to the disparities in data generation mechanisms between RCTs and RWDs, differences are likely and necessitate scrutiny to validate the merging of these datasets.

**Materials and Methods:** We compared the temporal and completeness characteristics of pharmaceutical RCT data from 5,734 diabetic kidney disease patients with corresponding RWD from electronic health records (EHRs) of 23,523 patients. Demographics, diagnoses, medications, laboratory measurements, and vital signs were analyzed using visualization, descriptive statistics, statistical testing, and cluster analysis.

**Results:** RCT and RWD sets exhibited significant differences in prevalence, longitudinality, completeness, and sampling density. The cluster analysis revealed distinct patient subgroups within both RCT and RWD sets, as well as clusters containing patients from both sets.

**Discussion and Conclusions:** The results highlight the differences between RCT and RWD datasets, and their respective data generation mechanisms. Nonetheless, in certain instances, RWD has the potential to enrich RCT data. These discrepancies should be taken into account during the planning stages of an RCT-RWD study, and we stress the importance of validation to verify the feasibility of combining RCT and RWD. Moreover, advanced methods are needed to mitigate these differences, for instance, when building an external control arm.

## BACKGROUND AND SIGNIFICANCE

Clinical real-world data (RWD) has garnered increasing interest for its use alongside clinical trial protocols in generating evidence in medical research. Several studies have explored the utilization of RWD in conducting clinical trials, [1, 2, 3, 4] providing external control groups for single-arm studies, [5, 6] and complementing the control group in randomized controlled trials (RCTs). [7, 8, 9] The rationale for using RWD to augment RCTs is premised on the assumption that RWD and RCT datasets are comparable despite potential biases. However, data generation mechanisms differ substantially between RWD and RCT, and systematic differences are probable. As a result, verifying the compatibility between datasets becomes a crucial component of data preprocessing to ensure the study’s feasibility.

Compared to clinical trial data, RWD’s quality can greatly fluctuate depending on its purpose and the specific dataset. Characteristics such as accuracy, completeness, and sampling intervals can vary among covariates, patients, and healthcare providers. [10, 11] Certain clinical measures, such as blood pressure and weight, are often available for research, but variables like outpatient medication exposure might be indirectly inferred from prescriptions and potentially overestimated if prescriptions were left unused. [11] Choices in study design, like index date or lookback window, can influence prevalence and incidence estimates based on claims and electronic health record (EHR) databases. [12] These factors may not be as clearly defined in RWD as in RCTs, potentially affecting the temporal alignment of the datasets. There might also be other underlying time-related biases which complicate, for instance, the identification of event onset or exposure to treatment. [13] For example, a delay between disease initiation and detection can introduce temporal variation and bias into the data. [13]

Apart from inclusion and exclusion criteria, the discrepancy between datasets can be alleviated through the application of appropriate validation criteria for the relevance, reliability, and quality of the RWD [14] and by particularly focusing on confounders related to exposure and outcome. [15] Several methods for selecting control patients have been suggested, such as cardinality matching of individuals [15] based on propensity scoring, [16] which has become the preferred method for adjusting group differences and reducing confounding. However, selection criteria and computational advancements are merely parts of the solution and should not be relied upon unconditionally.

In this study, we compared baseline data from a completed clinical trial on chronic kidney disease outcomes in individuals with type 2 diabetes to that from electronic health records (EHRs) in order to characterize their similarities and differences. Our focus was on five common data types: demographics, diagnoses, medications, laboratory measurements, and vital signs. We evaluated temporal aspects such as extent and sampling density, along with data completeness or missingness. Through statistical and cluster analysis, we demonstrate a partial overlap of RWD and RCT data, as well as their unique characteristics.

## METHODS AND MATERIALS

### Data

We used RCT data from the completed FIDELIO-DKD trial (Bayer, NCT02540993) to study the effect of finerenone on chronic kidney disease outcomes in type 2 diabetes in adult patients (≥ 18 years). [17] The data were pseudonymized, and we used only baseline data prior to randomization, including demographics, vital signs, diagnosis history, laboratory measurements, and concomitant medications. We obtained internal approval for secondary research use of the trial data.

To be included in the RWD set, patients needed to have chronic kidney disease as defined by ICD-10 codes N18, N19, or I12, or an estimated glomerular filtration rate (eGFR) <45 mL/min/1.73m2. Type 2 diabetes mellitus was also required, defined either by diagnosis code E11, the use of diabetes medication (ATC class A10), or a glycated hemoglobin (H-HbA1c) measurement ≥48 mmol/mol.

The RWD set contained data from patients who were first diagnosed with either chronic kidney disease or type 2 diabetes mellitus as adults. We assessed this post hoc using data from medications, diagnoses, and laboratory tests. We extracted these data along with demographics and vital signs from electronic healthcare records of HUS Helsinki University Hospital, Finland, covering a ten-year period from 2012 to 2021. The hospital approved the study (study permit HUS/230/2022).

The index date in the RCT data was the date of randomization, while in the RWD, it was the date when the chronic kidney disease inclusion criteria were met.

### Data structuring

The RCT data were structured. RWD were also structured, with the exception of smoking status, some medications, and New York Heart Association (NYHA) classes. We extracted these data from clinical documents using text mining and added them to the dataset with corresponding timestamps.

### Data harmonization

We used SNOMED coding to harmonize the different nomenclatures of RCT and RWD. The mapping was straightforward for demographics, medications, laboratory measurements, and vital signs. For diagnoses, 95.9 % of MedDRA codes in RCT and 94.1 % of ICD-10 codes in RWD were successfully mapped to SNOMED coding. Unmapped diagnoses codes, which were primarily in the Z-category indicating factors affecting health status or contact with health services, were not considered critical and were excluded from subsequent analyses. For diagnoses, we used the latest version of the mapping table from OHDSI Athena [18]. We performed standard unit conversions between RCT and RWD laboratory values for laboratory measurements.

### Analytics environment

We stored and processed the data on the HUS Acamedic cloud-based data analytics platform [19]. This platform enables high-performance scientific computing and can be scaled as necessary. The platform meets both European and national regulations (General Data Protection Regulation, Finlex 552/2019) for processing sensitive health and social data, it has a valid security certification, and is supervised by the National Supervisory Authority for Welfare and Health (Valvira).

### Statistical methods

We compared the prevalence of diagnoses and medications between RCT and RWD sets and reported counts and proportions by category. We compared the data longitudinality and sampling density aspects, namely the length of pre-index time, events per year, unique codes per patient, and interval between events, between RCT and RWD sets as continuous variables. These were reported with medians and interquartile ranges (IQR). We used chi-squared tests for group comparisons with categorical variables and the Kruskal-Wallis H test for continuous data. We performed the analyses with Python 3.8.10.0 using pandas (version 1.1.5) [20] and SciPy (version 1.5.3) [21] libraries.

### Cluster analysis of diagnoses and medications

To further examine and visually represent the differences and overlaps between the RCT and RWD sets, we performed separate cluster analyses for medication and diagnosis datasets, which were formed by merging the RCT and RWD sets. These analyses were similar to those outlined in [22]. For this purpose, we mapped diagnoses in the RCT data to International Classification of Diseases version 10 codes (ICD-10) as detailed in the Results section. Following the method in [22], we limited our analysis to ICD-10 diagnosis categories A to N with a precision of three characters. Hence, our focus was on disorders while excluding codes related to pregnancy, external causes, malformations, and contacts with health services. For medication data, we selected the first four characters of the Anatomical Therapeutical Chemical (ATC) codes. In both data sets, a specific code had to have at least 1% prevalence to be included in the analysis. Consequently, the data sets contained 65 covariates for diagnoses and 84 covariates for medications.

Following a two-step approach for clustering [22], we initially trained a variational autoencoder (VAE) model [23, 24] using Keras [25] (version 2.3.1). This model projected binary diagnosis vectors into a two-dimensional latent space [23, 24]. Then we clustered the projected vectors using the HDBSCAN algorithm [26].

For the encoder and decoder components of the VAE models, we utilized fully connected multilayer perceptrons (MLPs) with a single hidden layer, with either a hyperbolic tangent (tanh) or a rectified linear unit (ReLU) [27] as the activation function. Since the purpose of the cluster analysis was to visually distinguish the data sets and their differences, we selected the activation function that produced visually distinct subgroups. The VAE model was trained using the evidence lower bound objective, which comprises a reconstruction loss and a regularizer on the latent space. We divided the data into a training set (90% of the data) and a validation set (10% of the data), with the validation data used to choose a hyperparameter for the number of gradient descent steps for training. Eventually, we mapped both sets to the latent space for the clustering step.

We used the HDBSCAN algorithm [26] (hdbscan version 0.8.29) to extract clusters for subsequent description and interpretation of the identified subgroups. For diagnosis data, we used the HDBSCAN parameters min_cluster_size=220 and min_samples=1, and for medications data, we used min_cluster_size=200 and min_samples=5. To visualize the results of the cluster analysis, we used NumPy [28] (version 1.21.6) to compute two-dimensional histograms, SciPy [21] (version 1.5.3) for kernel density estimation, and Matplotlib [29] (version 3.2.1) for plotting. To preserve patient anonymity, we avoided presenting the locations of individual data points and instead utilized histogram and density-based approaches for visualization.

## RESULTS

### Qualitative comparisons

We analyzed the pre-index baseline data of 23,523 RWD and 5,734 RCT patients, all of whom had both chronic kidney disease and type 2 diabetes. Harmonizing to common nomenclature for both RCT and RWD sets was straightforward, but we noted considerable qualitative differences in data generation, temporality, and completeness, as summarized in Table 1. For instance, in the RCT data, diagnoses and concomitant medications based on case report forms (CRFs) collected by investigators spanned up to 50 years pre-index. Conversely, in the RWD, all diagnoses in EHRs with precise dates were available up to 10 years pre-index, constrained by the research permit. Laboratory measurements and vital signs were only available near index in RCT data and up to 10 years pre-index in RWD.

**Table 1:** Qualitative observations between the five medical domains in the RCT and RWD data sets.

| Property \ Data | Demographics (DM) | Diagnosis history (DH) | Laboratory measurements (LB) | Vital signs (VS) | Concomitant medications (CM) |
| --- | --- | --- | --- | --- | --- |
| **Data generation** | RCT: Measured from patients at screening.<br>RWD: As measured in healthcare. | RCT: CRF and discretion of a physician.<br>RWD: All diagnoses during the years as in an institutional database. | RCT: Measured from patients at screening.<br>RWD: As measured in healthcare. | RCT: Measured from patients at screening.<br>RWD: As measured in healthcare. | RCT: CRF and discretion of a physician.<br>RWD: Prescriptions and inpatient drugs administered. Mentions in text. |
| **Longitudinality** | RCT: Values available at index date (time of randomization).<br>RWD: Time series. Index date was defined to calculate age and other variables at index. | RCT: Time series, up to 50 years of historical data. No exact dates.<br>RWD: Time series up to 10 years of historical data, limited by research permit. Exact dates. | RCT: Only available near index date.<br>RWD: Time series up to 10 years of historical data available, limited by research permit. Exact dates. | RCT: Only available near index date.<br>RWD: Time series up to 10 years of historical data available, limited by research permit. Exact dates. | RCT: Time series, up to 50 years of historical data. No exact dates.<br>RWD: Time series up to 10 years of historical data, limited by research permit. Exact dates. |
| **Completeness** | RCT: Values available at index date.<br>RWD: Obtained from time series of structured data except smoking status and NYHA class which were extracted from structured and text sources. | RCT: Some diagnoses underrepresented, such as infections (Figure 1).<br>RWD: Some diagnoses underrepresented, such as genitourinary diseases (Figure 1). | RCT: All values available from a given set of measurements.<br>RWD: Missing values with respect to trial design, availability depends on healthcare practices. | RCT: All values available from a given set of measurements.<br>RWD: Missing values with respect to trial design, availability depends on healthcare practices. | RCT: Selected medications underrepresented in the data, such as anti-infectives (Figure 1).<br>RWD: All prescriptions and administered inpatient medications in institutional database. Selected medications extracted from text. |
| **Nomenclature** | Commonly used demographic variables, such as age, gender, and smoking. | RCT: MedDRA codes.<br>RWD: ICD-10 codes. | RCT: MedDRA codes.<br>RWD: Logical Observation Identifiers Names and Codes (LOINC). | Commonly used vital sign variables, such as BMI, blood pressure and heart rate. | ATC codes both in RCT and RWD structured data. Also, medication labels in text. |
| **Harmonization** | Same nomenclature | RCT: 95.9 % of MedDRA codes successfully mapped to SNOMED.<br>RWD: 94.1 % of ICD-10 codes successfully mapped to SNOMED. | Straightforward mapping between laboratory measurements. Standard unit conversions were performed between RCT and RWD laboratory values. | Same nomenclature | Same nomenclature |

### Quantitative comparisons

Statistical analysis revealed notable differences in completeness, particularly in medications and diagnoses. After harmonizing the different nomenclatures of RCT and RWD, we used ATC and ICD-10 code classes for easier interpretation of the results. In concomitant medications (Figure 1A), the most significant difference in prevalence was observed in anti-infectives for systemic use (class J, RCT 8.6%, RWD 66.0 %, P<0.001). In diagnosis history (Figure 1B), the largest difference in prevalence was seen in class R, which refers to symptoms, signs, and abnormal clinical and laboratory findings (RCT 14.3%, RWD 57.1%, P<0.001). Although fulfilling the inclusion criteria, not all patients in the RWD had both inclusion diagnoses of type 2 diabetes and chronic kidney disease, which belong to classes E (endocrine, nutritional, and metabolic diseases, RCT 100.0%, RWD 50.7%, P<0.001) and N (diseases of the genitourinary system, RCT 100.0%, RWD 49.9%, P<0.001), respectively.

**Figure 1.**
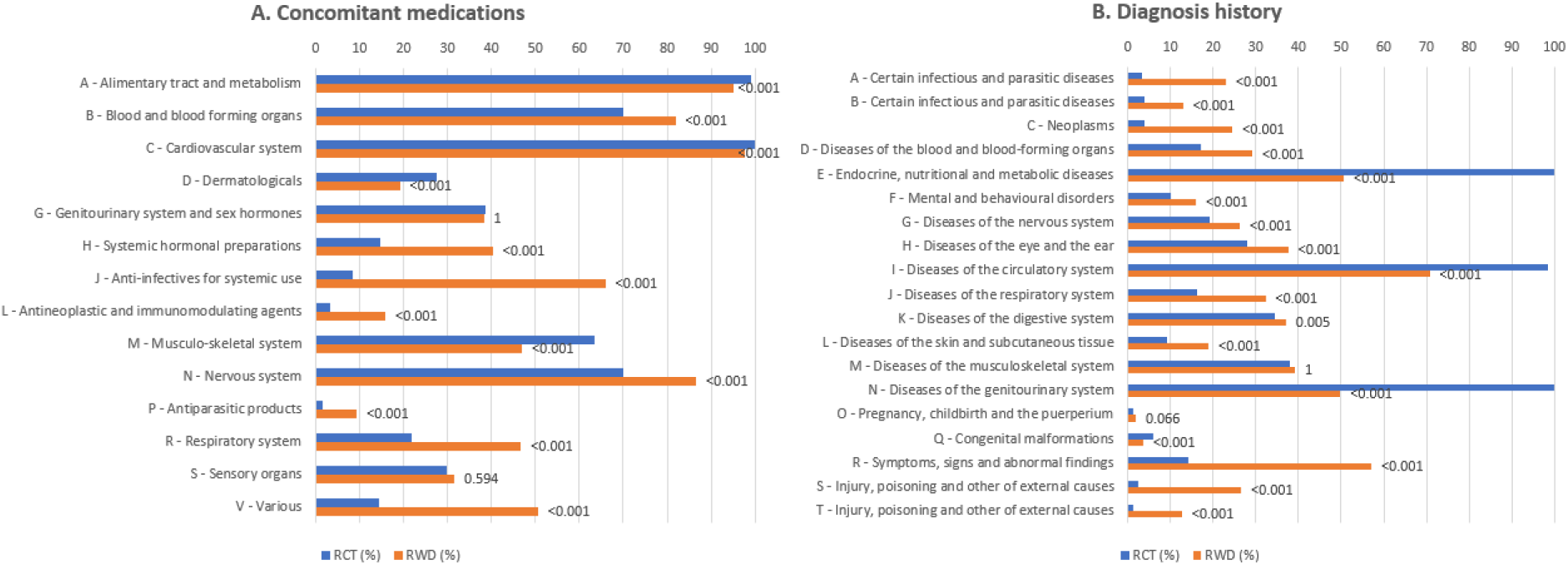
Comparison of proportion (%) of RCT (N = 5 734) and RWD (N = 23 523) patients with different medication/diagnosis types recorded, each pair of RCT and RWD bars corresponding to an ATC class (A) or ICD-10 class (B). Value labels above bars correspond to P values calculated with chi-squared tests. Presented P values were adjusted with Bonferroni correction to address the issue of multiple comparisons.

As demographics, laboratory measurements, and vital signs in the RCT data were only available near the index time, we further analyzed the temporal characteristics of concomitant medications and diagnoses (Table 2). The pre-index time (in years) from the first event to the index date was significantly longer in RCT data for both medications (median 7.7 vs. 4.9, P<0.001) and diagnoses (median 20.3 vs. 5.2, P<0.001). We also assessed the time interval between events and the sampling density, defined as the number of events per year. The number of events per year and the number of unique codes were significantly larger in RWD compared to trial data for both medications and diagnoses. Consequently, the time interval between consecutive events was significantly shorter in RWD for both medications and diagnoses compared to RCT. Additionally, we observed a significant difference between RWD and RCT cohorts in age at index (median 76 years in RWD, median 66 in RCT, P<0.001) and sex male/female ratio (RWD 47/53, RCT 70/30, P<0.001).

**Table 2:** Comparison of longitudinality and sampling density variables between concomitant medication and diagnosis domains for the RCT (N = 5 734) and RWD (N = 23 523) patients.

| Concomitant medications |  |  |  |  | Diagnosis history |  |  |  |  |
| --- | --- | --- | --- | --- | --- | --- | --- | --- | --- |
| Variable | Quartile | RCT | RWD | P value* | Variable | Quartile | RCT | RWD | P value* |
| Pre-index time (Years) | 25 % | 3,6 | 2,4 | <0.001 | Pre-index time (Years) | 25 % | 14,9 | 2,7 | <0.001 |
|  | Median | 7,7 | 4,9 |  |  | Median | 20,3 | 5,2 |  |
|  | 75 % | 13,7 | 6,5 |  |  | 75 % | 28,7 | 7,3 |  |
| Number of events | 25 % | 4 | 9 | <0.001 | Number of events | 25 % | 4 | 5 | <0.001 |
|  | Median | 6 | 20 |  |  | Median | 5 | 13 |  |
|  | 75 % | 9 | 38 |  |  | 75 % | 7 | 25 |  |
| Number of events per year | 25 % | 0,5 | 2,9 | <0.001 | Number of events per year | 25 % | 0,2 | 1,7 | <0.001 |
|  | Median | 0,8 | 5,4 |  |  | Median | 0,3 | 3,3 |  |
|  | 75 % | 1,6 | 10,0 |  |  | 75 % | 0,4 | 7,6 |  |
| Number of unique medications | 25 % | 10 | 14 | <0.001 | Number of unique diagnoses | 25 % | 5 | 5 | <0.001 |
|  | Median | 14 | 23 |  |  | Median | 7 | 9 |  |
|  | 75 % | 19 | 34 |  |  | 75 % | 9 | 15 |  |
| Interval between events (Days) | 25 % | 113,3 | 3,0 | <0.001 | Interval between events (Days) | 25 % | 464,6 | 13,0 | <0.001 |
|  | Median | 288,0 | 13,5 |  |  | Median | 914,0 | 31,0 |  |
|  | 75 % | 576,5 | 41,0 |  |  | 75 % | 1826,0 | 89,0 |  |

\* Presented P values were adjusted with Bonferroni correction to address the issue of multiple comparisons.

### Cluster analysis of diagnoses and medications

To elucidate the overlap between RWD and RCT cohorts, we conducted cluster analyses of diagnoses and medications, integrating the RWD and RCT datasets for this purpose. Figures 2 and 3 graphically represent the outcomes of these analyses. In both the diagnosis and medication datasets, the RWD and RCT cohorts emerged as discernable subgroups in the latent space of the Variational Autoencoder (VAE) model. Eleven clusters were discerned from the diagnosis data, and seven from the medication data. For the diagnosis dataset, 5133 patients (18%) did not align with any cluster; a comparable figure for the medication dataset was 5487 patients (19%). Detailed overviews of each cluster can be found in Tables 3 and 4, while Supplementary files 1 and 2 provide prevalence data for each diagnosis and medication within each cluster.

**Table 3:**
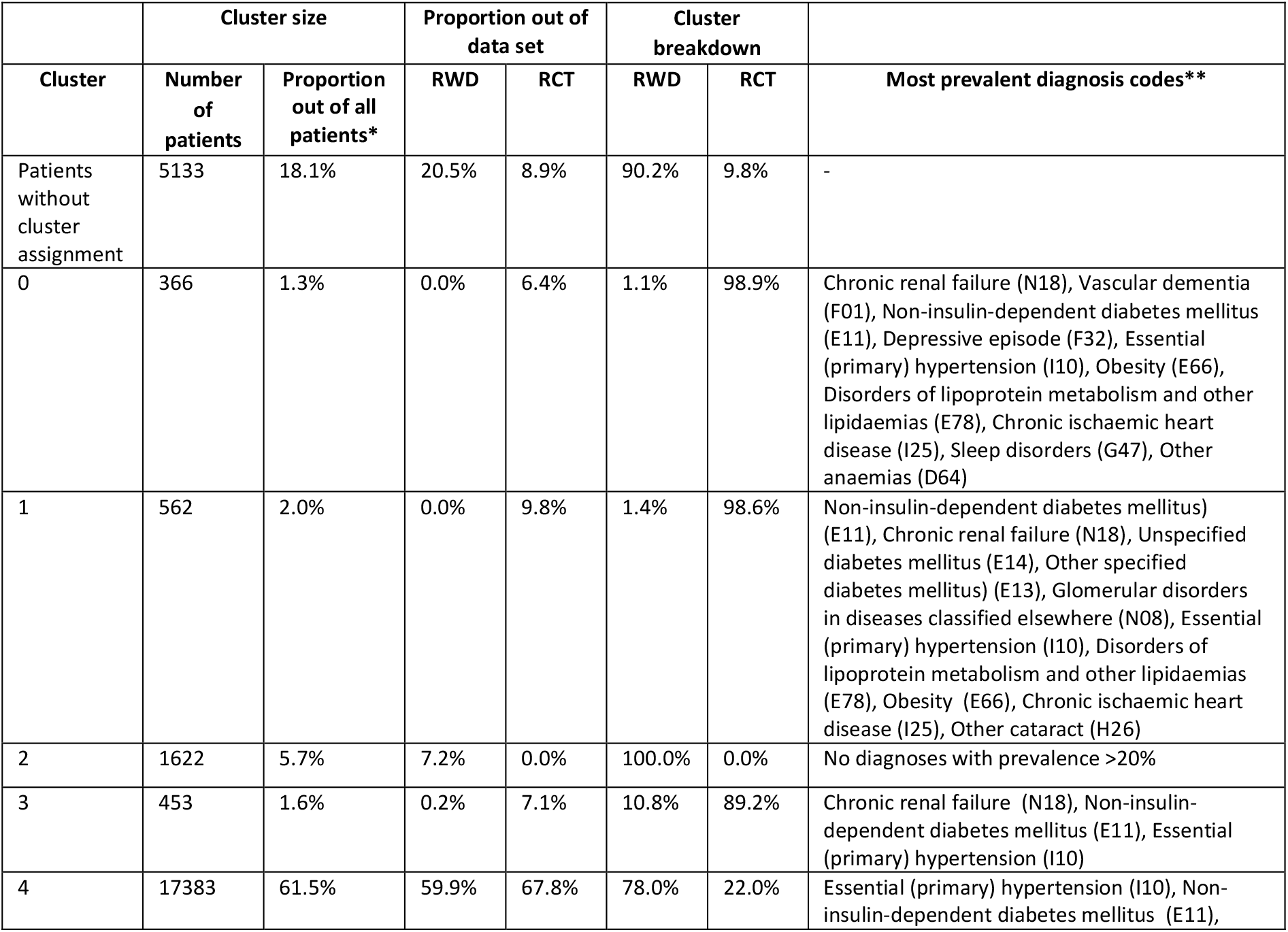
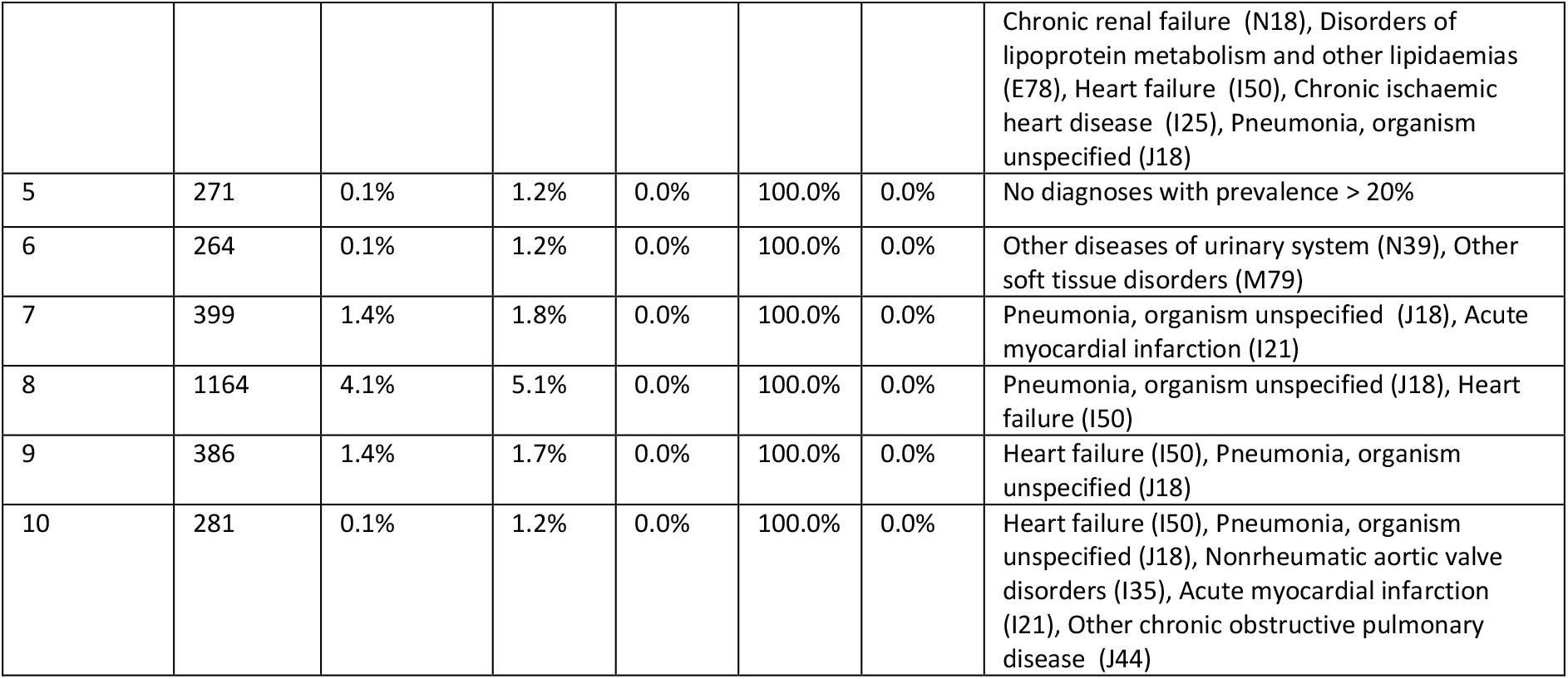
Results from clustering of diagnosis data. Proportion out of data set describes how the RWD and RCT patients are distributed along the different clusters. Cluster breakdown shows how large proportions of the patients belonging to a cluster come from RWD and RCT sets. *All patients used for the clustering of diagnosis data set. **Diagnoses with at least 20% prevalence in the cluster are listed in descending order of prevalence out of 65 diagnosis codes used for VAE model training.

**Table 4:** Results from clustering of medications data. Proportion out of data set describes how the RWD and RCT patients are distributed along the different clusters. Cluster breakdown shows how large proportions of the patients belonging to a cluster come from RWD and RCT sets. *All patients used for the clustering of medication data set. **Medications with at least 60% prevalence in the cluster are presented in descending order of prevalence out of 84 medication codes used for VAE model training.

| Cluster | Cluster size |  | Proportion out of data set |  | Cluster breakdown |  | Most prevalent medication codes** |
| --- | --- | --- | --- | --- | --- | --- | --- |
|  | Number of patients | Proportion out of all patients* | RWD | RCT | RWD | RCT |  |
| Patients without cluster assignment | 5487 | 19.4% | 17.4% | 27.5% | 71.6% | 28.4% | - |
| 0 | 630 | 2.2% | 0.3% | 9.9% | 11.0% | 89.0% | Ace inhibitors, plain (C09A), Blood glucose lowering drugs, excl. insulins (A10B), Lipid modifying agents, plain (C10A) |
| 1 | 453 | 1.6% | 0.0% | 8.0% | 0.0% | 100.0% | Stomatological preparations (A01A), Topical products for joint and muscular pain (M02A), Antithrombotic agents (B01A), ACE inhibitors, plain (C09A), Other analgesics and antipyretics (N02B), Lipid modifying agents, plain (C10A), Blood glucose lowering drugs, excl. insulins (A10B), Insulins and analogues (A10A), Beta blocking agents (C07A) |
| 2 | 1674 | 5.9% | 7.4% | 0.2% | 99.5% | 0.5% | Other antianemic preparations (B03X), Other analgesics and antipyretics (N02B), Antithrombotic agents (B01A), Opioids (N02A), High-ceiling diuretics (C03C), Drugs for peptic ulcer and gastro-oesophageal reflux disease (GORD) (A02B), Calcium (A12A), Insulins and analogues (A10A), Selective calcium channel blockers with mainly vascular effects (C08C), Beta blocking agents (C07A), Drugs for constipation (A06A), Lipid modifying agents, plain (C10A), Other beta-lactam antibacterials (J01D), Vitamin A and D, incl. combinations of the two (A11C), Antiemetics and antinauseants (A04A), Corticosteroids for systemic use, plain (H02A), Iron preparations (B03A), Hypnotics and sedatives (N05C), Anxiolytics (N05B), Beta-lactam antibacterials, penicillins (J01C) |
| 3 | 875 | 3.1% | 0.1% | 15.1% | 2.6% | 97.4% | Angiotensin II receptor blockers (ARBs), plain (C09C), Blood glucose lowering drugs, excl. insulins (A10B), Lipid modifying agents, plain (C10A), Selective calcium channel blockers with mainly vascular effects (C08C), Insulins and analogues (A10A) |
| 4 | 17751 | 62.9% | 74.8% | 15.1% | 95.2% | 4.8% | Antithrombotic agents (B01A), Other analgesics and antipyretics (N02B), Beta blocking agents (C07A), Opioids (N02A), Lipid modifying agents, plain (C10A), High-ceiling diuretics (C03C), Drugs for peptic ulcer and gastro-oesophageal reflux disease (GORD) (A02B), Other beta-lactam antibacterials (J01D), Insulins and analogues (A10A), Blood glucose lowering drugs, excl. insulins (A10B), Hypnotics and sedatives (N05C), Drugs for constipation (A06A), Selective calcium channel blockers with mainly vascular effects (C08C) |
| 5 | 937 | 3.3% | 0.0% | 16.6% | 0.0% | 100.0% | Stomatological preparations (A01A), Antithrombotic agents (B01A), Other analgesics and antipyretics (N02B), Topical products for joint and muscular pain (M02A), Angiotensin II receptor blockers (ARBs), plain (C09C), Lipid modifying agents, plain (C10A), Blood glucose lowering drugs, excl. insulins (A10B), Selective calcium channel blockers with mainly vascular effects (C08C), Insulins and analogues (A10A) |
| 6 | 436 | 1.5% | 0.0% | 7.7% | 0.7% | 99.3% | Stomatological preparations (A01A), Lipid modifying agents, plain (C10A), Antithrombotic agents (B01A), Topical products for joint and muscular pain (M02A), Other analgesics and antipyretics (N02B), Blood glucose lowering drugs, excl. insulins (A10B), Selective calcium channel blockers with mainly vascular effects (C08C), Insulins and analogues (A10A), Angiotensin II receptor blockers (ARBs), plain (C09C) |

**Figure 2.**
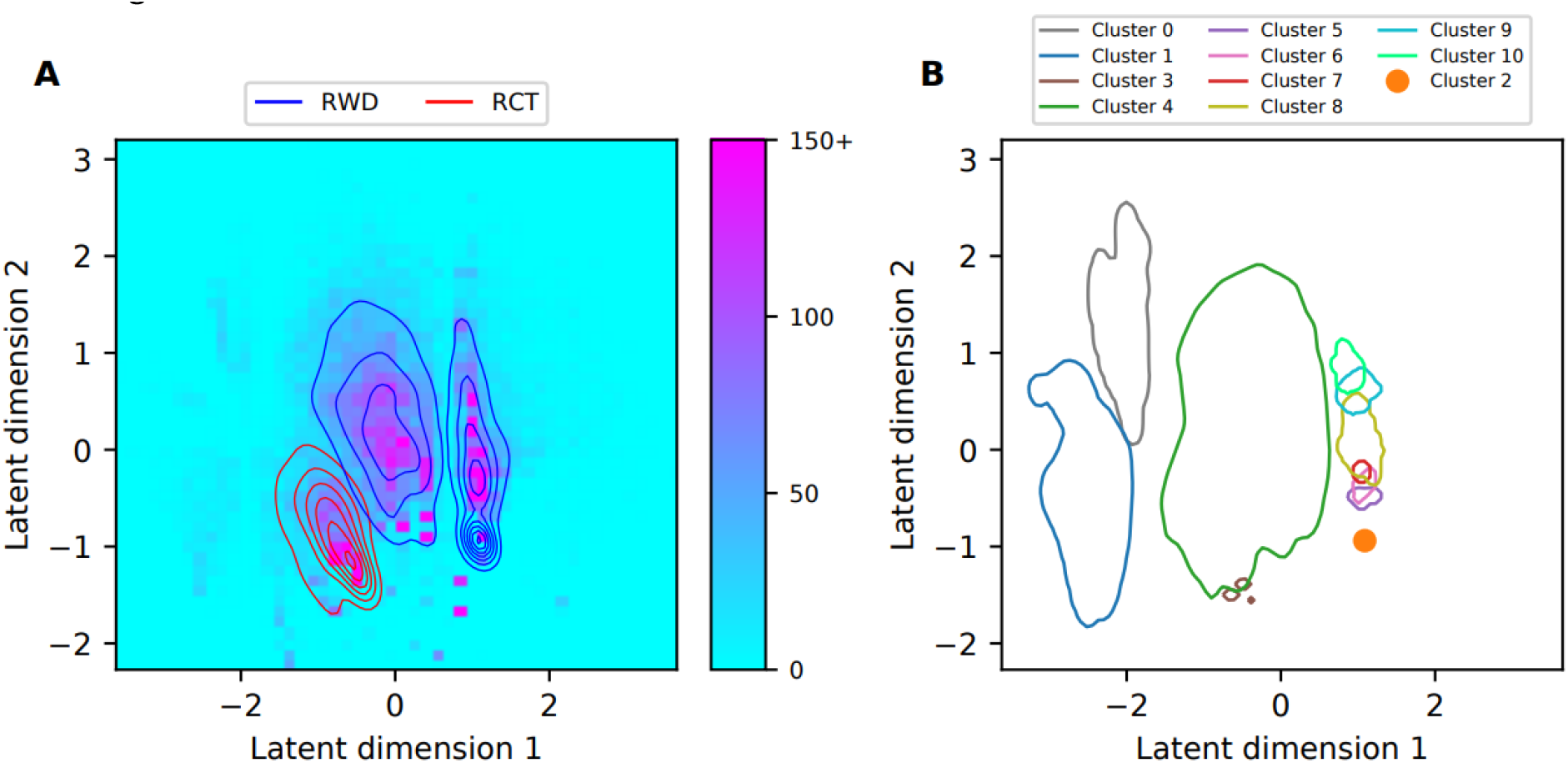
Visualization of the results from the cluster analysis of the diagnosis data. (A) The two-dimensional histogram shows the density of the combined RCT and RWD data sets along the two dimensions of the learned latent representation. We truncated histogram bin counts to 150. The red and blue lines show the Gaussian kernel density estimates (KDE) of the RCT and RWD distributions, respectively. (B) Contour lines for the Gaussian KDEs fitted using the datapoints belonging to clusters 0 and 3–10, plotted at the 0.05 value of each probability density estimate. The datapoints in cluster 2 lie very near to each other, leading to kernel density estimation failing, which is why we visualized the mean of these data point locations instead.

**Figure 3.**
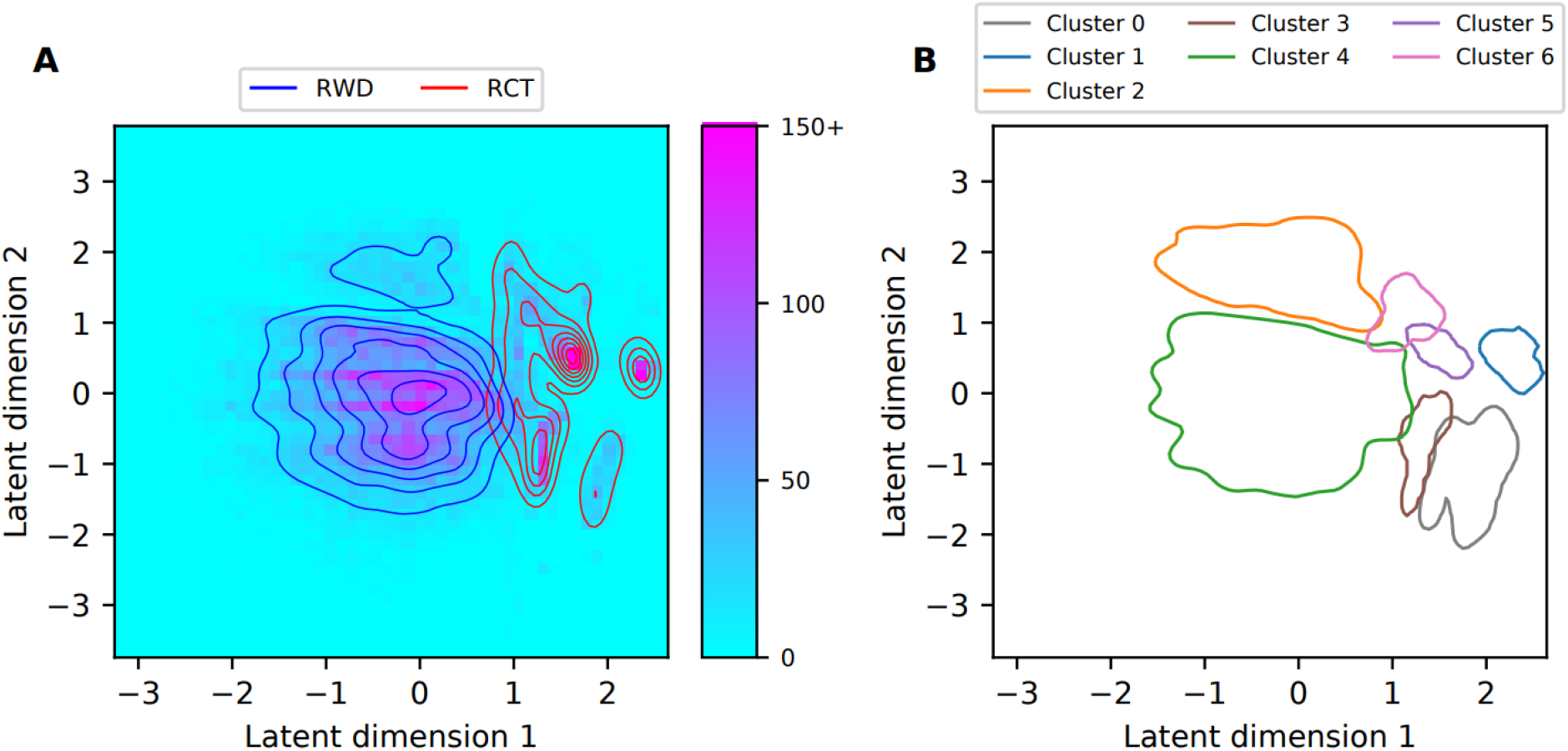
Visualization of the results from the cluster analysis of the medications data. (A) The two-dimensional histogram shows the density of the combined RCT and RWD data sets along the two dimensions of the learned latent representation. We truncated histogram bin counts to 150. The red and blue lines show the Gaussian kernel density estimates (KDE) of the RCT and RWD distributions, respectively. (B) Contour lines for the Gaussian KDEs fitted using the datapoints belonging to each cluster, plotted at the 0.05 value of each probability density estimate.

From the diagnosis dataset, three clusters (0, 1, and 3) mainly consisted of RCT patients, while eight clusters (2, 4-10) primarily consisted of RWD patients. Cluster 4 demonstrated the most substantial overlap, with 22% of its patients from the RCT cohort and 78% from the RWD cohort. Cluster 3 also exhibited significant overlap, with 89% of its data derived from the RCT cohort and 11% from the RWD cohort. This overlap signifies the presence of both RCT and RWD patients within a single cluster. Importantly, over half of both the RCT and RWD cohorts were consolidated within Cluster 4, whereas the proportions for other clusters were substantially lower. Clusters 2 and 5 were distinguished by a very low prevalence for all diagnoses utilized in VAE model training.

Aside from these two clusters, those predominantly composed of RWD patients could be loosely divided into two groups: the larger Cluster 4 containing 17383 patients, and a group comprising the adjacent Clusters 6-10. In Cluster 4, essential hypertension (I10) and non-insulin-dependent diabetes (E11) were the most prevalent diagnoses. Clusters 7-10 were characterized by prevalent diagnoses of pneumonia (J18) and either heart failure (I150), acute myocardial infarction (I21), or both. Cluster 6 differed substantially from Clusters 7-10, although geographically close. The most common diagnoses for Cluster 6 were other diseases of the urinary system (N39) and other soft tissue disorders (M79).

The RCT-dominated clusters (0, 1, and 3) shared prevalent diagnosis codes for chronic renal failure (N18), non-insulin-dependent diabetes (E11), and essential hypertension (I10). Additional prevalent diagnoses in Cluster 0, but not in Cluster 1, included vascular dementia (F01) and depressive episode (F32). Cluster 1, but not Cluster 0, exhibited a higher prevalence of other specified diabetes mellitus (E13), unspecified diabetes mellitus (E14), and glomerular disorders in diseases classified elsewhere (N08).

From the medication dataset, two clusters (2 and 4) were mostly composed of RWD patients, and five (0, 1, 3, 5, and 6) were largely comprised of RCT patients. The most substantial overlap between RCT and RWD cohorts was observed in Cluster 0 (11% RWD, 89% RCT) and the largest Cluster 4 (5% RCT, 95% RWD). Over 70% of the RWD patients were included in Cluster 4, and the proportions for the remaining clusters were significantly smaller.

Conversely, RCT patients were more evenly distributed across the clusters Clusters 3, 4, and 5 each comprised approximately 15% of the RCT patients, with the remaining clusters containing smaller proportions. Across all clusters, the most frequently prescribed medications (prevalence >60%) incorporated at least one medication code pertaining to the cardiovascular system (ATC codes beginning with ‘C’) and at least one code related to diabetes medications (ATC codes beginning with ‘A*’).

Contrary to other clusters, neither antithrombotic agents (B01A) nor opioids (N02A) and other analgesics and antipyretics (N02B) were prevalent in Clusters 0 and 3. Clusters 1, 5, and 6 all shared stomatological preparations (A01A) as their most frequent medication code, whereas this code was scarcely seen in Clusters 2 and 4. Clusters 2 and 4 were distinctive in that they had high prevalences of other beta-lactam antibacterials (J01D) and hypnotics and sedatives (N05C). In addition, other antianemic preparations (B03X) and calcium (A12A) were among the medications specific to Cluster 2.

## DISCUSSION

This study illustrates that real-world data (RWD) and randomized controlled trial (RCT) datasets, derived from patients with diabetic chronic kidney disease, share common characteristics but also exhibit substantial differences in terms of data generation, completeness, and temporal dynamics. These discrepancies have implications for study design validity and mandate careful examination when merging RWD and RCT data.

### Data generation and longitudinality

The noted differences between RCT and RWD predominantly arise from their respective data generation processes and objectives. RCT data is prospectively collected following a specified study protocol, whereas RWD is extracted through queries from hospital data infrastructure. For instance, in RCT data, only the initial diagnosis date is recorded by the investigator on case report forms (CRFs), potentially leading to selection and recall bias. Conversely, in RWD, each inpatient and outpatient diagnosis are precisely dated; however, data from a single institution may not encompass the patient’s entire medical history. Unlike RWD, RCT data aims to assess the efficacy and safety of an intervention. Therefore, data elements unrelated to the trial’s exposure and outcome, such as anti-infective medications and symptom diagnoses in our study, may be underrepresented.

Furthermore, specific elements of RCT data, such as laboratory measurements, may only be cross-sectional and timed near the index date. Conversely, electronic health record (EHR) data chronicles patient interaction with healthcare services and inherently contains longitudinal data without defined start or end dates, possibly spanning decades. In our study, our research permit limited RWD to a ten-year range. Thus, incomplete patient history, whether derived from CRFs or EHRs, introduces biases and differences between RCT and RWD data. Ideally, RCT baseline data should utilize RWD covering the complete patient history over the relevant time range.

### Data density and completeness

Diagnoses and medications were sampled significantly more densely in our RWD set compared to RCT. Consequently, RWD offered a more accurate portrayal of the patients’ state by capturing all pertinent data. Nevertheless, data completeness and accuracy can be limited if data is sourced solely from a single healthcare provider. Despite all patients meeting the inclusion criteria, some lacked records of chronic kidney disease or type 2 diabetes diagnosis in RWD data, unlike in the RCT data.

These diagnoses may be partly recorded in primary healthcare data, which was not included in this study. Additionally, text mining was necessary to extract data from unstructured texts in RWD. Despite these limitations, our findings suggest that, in certain scenarios, RWD could supplement RCT data through EHR to electronic data capture (EHR2EDC) automation. [30]

The harmonization process resulted in a minor loss of diagnoses, with 94.1% of RWD and 95.9% of RCT diagnosis codes successfully mapped to SNOMED codes. The mapping process for the remaining data types was straightforward.

### Cluster analysis

We utilized cluster analysis on the combined real-world and randomized controlled trial (RCT) datasets to illustrate the heterogeneity of the study population, discover patient subgroups, and assess the overlap between the two datasets. Our analysis revealed that the datasets were largely distinct. Both RCT and real-world data (RWD) sets comprised unique subgroups, with an overlap observed in only a few clusters. This was true for both diagnosis and medication datasets. However, in the diagnosis data, the largest cluster (cluster 4) contained a significant number of patients from both RWD and RCT sets. Hence, even though overlap was not present in all clusters, many RWD and RCT patients were grouped together in the cluster analysis.

The cluster analyses underscore the challenges in finding overlaps between real-world and RCT data, emphasizing the necessity for advanced methods to identify matching external controls. In the cluster analyses, we chose input covariates using a prevalence threshold of 1%, leading to 65 and 84 input covariates for diagnoses and medications data, respectively. A higher threshold would yield fewer covariates that could potentially differentiate the real-world and RCT datasets. The covariate set chosen for aligning RCT and RWD can influence the outcome and therefore requires careful selection. In addition to clinical differences between groups, there could be disparities in data completeness, i.e., how well the selected covariates are captured in the different data sources. It’s important to note that the clustering results are influenced by our choices made in the VAE model training and cluster analysis and represent one set of possible options. Furthermore, we conducted the cluster analyses without considering possible demographic differences between RCT and RWD, which could account for some of the identified differences. However, cluster analysis proved beneficial in identifying patients with overlapping characteristics in RCT and RWD.

Our results indicate the overlap and discrepancies in one trial and RWD pair but cannot be directly generalized. Nevertheless, similar observations are likely in any study that merges RCT and RWD sources.

## CONCLUSION

In conclusion, RCT and RWD exhibit substantial differences in data longitudinality, completeness, sampling density, among other factors, all of which should be considered when designing studies that amalgamate data from these sources. Despite their inherent limitations, RWD sources could, in certain instances, be used to enrich RCT datasets, for instance, to enhance the longitudinality and completeness of patient history. RCT and RWD sets were distinct and could form unique patient subgroups, which must be taken into account in studies merging RCT and RWD and in patient matching.

## Data Availability

Regarding the RWD, according to the Finnish legislation, access to individual-level data is restricted only to individuals named in the study permit. The study protocol is available upon request from the corresponding author. Regarding the RCT data, the data are not publicly available due to containing information that could compromise research participant privacy/consent.

## AUTHOR CONTRIBUTIONS

Conception of the work: Miika Koskinen, Jussi Leinonen

Study design: Samu Kurki, Viivi Halla-aho, Jussi Leinonen, Miika Koskinen

Acquisition of data: Samu Kurki, Viivi Halla-aho, Jussi Leinonen, Miika Koskinen

Analysis: Samu Kurki, Viivi Halla-aho

Interpretation: All authors

Drafting the manuscript text: Samu Kurki, Viivi Halla-aho, Jussi Leinonen, Miika Koskinen

Revision of the manuscript critically for important intellectual content: All authors

Final approval of the version to be published: All authors

Agreement to be accountable for all aspects of the work in ensuring that questions related to the accuracy or integrity of any part of the work are appropriately investigated and resolved: All authors

## SUPPLEMENTARY MATERIAL

Supplementary file 1: Clustering results tables for diagnosis data set

Supplementary file 2: Clustering results tables for medication data set

## ACKNOWLEDGEMENTS

The authors would like to acknowledge valuable support by Senior Statistician Kaisa Laapas, and Senior Statistician, PhD Alberto Pessia at Bayer Oy. The computation of the results presented in this work has been performed using the resources at HUS Acamedic analytics platform.

## REFERENCES

[1] D. Abrahami, R. Pradhan, H. Yin, P. Honig, E. B. Andre and L. Azoulay, “Use of Real-World Data to Emulate a Clinical Trial and Support Regulatory Decision Making: Assessing the Impact of Temporality, Comparator Choice, and Method of Adjustment,” Clinical Pharmacology & Therapeutics, vol. 109, no. 2, pp. 452–461, 2021.

[2] T. Jemielita, X. Li, B. Piperdi, W. Zhou, T. Burke and C. Chen, “Overall survival with second-line pembrolizumab in patients with non--small-cell lung cancer: Randomized phase III clinical trial versus propensity-adjusted real-world data,” JCO clinical cancer informatics, no. 5, pp. 56–65, 2021.

[3] Z. Chen, H. Zhang, Y. Guo, T. J. George, M. Prosperi, W. R. Hogan, Z. He, E. A. Shenkman, F. Wang and J. Bian, “Exploring the feasibility of using real-world data from a large Clinical Data Research Network to simulate clinical trials of Alzheimer’s disease,” NPJ digital medicine, vol. 4, no. 1, pp. 1–9, 2021.

[4] J. R. Rogers, J. Lee, Z. Zhou, Y. K. Cheung, G. Hripcsak and C. Weng, “Contemporary use of real-world data for clinical trial conduct in the United States: a scoping review,” Journal of the American Medical Informatics Association, vol. 28, no. 1, pp. 144–154, 2021.

[5] G. Carrigan, S. Whipple, W. B. Capra, M. D. Taylor, J. S. Brown, M. Lu, B. Arnieri, R. Copping and K. J. Rothman, “Using electronic health records to derive control arms for early phase single-arm lung cancer trials: proof-of-concept in randomized controlled trials,” Clinical Pharmacology & Therapeutics, vol. 107, no. 2, pp. 369–377, 2020.

[6] J. Davies, M. Martinec, P. Delmar, M. Coudert, W. Bordogna, S. Golding, R. Martina and G. Crane, “Comparative effectiveness from a single-arm trial and real-world data: alectinib versus ceritinib,” Journal of comparative effectiveness research, vol. 7, no. 9, pp. 855–865, 2018.

[7] T. Jemielita, L. Widman, C. Fox, S. Salomonsson, K.-L. Liaw and A. Pettersson, “Replication of Oncology Randomized Trial Results using Swedish Registry Real World-Data: A Feasibility Study,” Clinical Pharmacology & Therapeutics, vol. 110, no. 6, pp. 1613–1621, 2021.

[8] C. Patry, L. D. Sauer, A. Sander, K. Krupka, A. Fichtner, J. Brezinski, Y. Geissbühler, E. Aubrun, A. Grinienko, L. D. Strologo, D. Haffner, J. Oh, R. Gernda, L. Pape, R. Topaloglu, L. T. Weber, A. Bouts, J. J. Kim, A. Prytula, J. König, M. Shenoy, B. Höcker and B. Tönshoff, “Emulation of the control cohort of a randomized controlled trial in pediatric kidney transplantation with Real-World Data from the CERTAIN Registry,” Pediatric Nephrology, pp. 1–12, 2022.

[9] S. Popat, S. V. Liu, N. Scheuer, G. G. Hsu, A. Lockhart, S. V. Ramagopalan, F. Griesinger and V. Subbiah, “Addressing challenges with real-world synthetic control arms to demonstrate the comparative effectiveness of Pralsetinib in non-small cell lung cancer,” Nature communications, vol. 13, no. 1, pp. 1–10, 2022.

[10] K. J. Lin, R. J. Glynn, D. E. Singer, S. N. Murphy, J. Lii and S. Schneeweiss, “Out-of-system care and recording of patient characteristics critical for comparative effectiveness research,” Epidemiology, vol. 29, no. 3, pp. 356–363, 2018.

[11] J. M. Franklin and S. Schneeweiss, “When and how can real world data analyses substitute for randomized controlled trials?,” Clinical Pharmacology & Therapeutics, vol. 102, no. 6, pp. 924–933, 2017.

[12] J. A. Rassen, D. Bartels, S. Schneeweiss, A. R. Patrick and W. Murk, “Measuring prevalence and incidence of chronic conditions in claims and electronic health record databases,” Clinical epidemiology, pp. 1–15, 2019.

[13] S. Suissa and S. Dell’Aniello, “Time-related biases in pharmacoepidemiology,” Pharmacoepidemiology and drug safety, vol. 29, no. 9, pp. 1101–1110, 2020.

[14] M. Ghadessi, R. Tang, J. Zhou, R. Liu, C. Wang, K. Toyoizumi, C. Mei, L. Zhang, C. Deng and R. A. Beckman, “A roadmap to using historical controls in clinical trials--by Drug Information Association Adaptive Design Scientific Working Group (DIA-ADSWG),” Orphanet Journal of Rare Diseases, vol. 15, no. 1, pp. 1–19, 2020.

[15] B. Nikram and J. Zubizarreta, “Using cardinality matching to design balanced and representative samples for observational studies,” JAMA, vol. 327, no. 2, pp. 173–174, 2022.

[16] P. R. Rosenbaum and D. B. Rubin, “The central role of the propensity score in observational studies for causal effects,” Biometrika, vol. 70, no. 1, pp. 41–55, 1983.

[17] G. Bakris, R. Agarwal, S. Anker, B. Pitt, L. Ruilope, P. Rossing, P. Kolkhof, C. Nowack, P. Schloemer, A. Joseph and G. Filippatos, “Effect of Finerenone on Chronic Kidney Disease Outcomes in Type 2 Diabetes,” New England Journal of Medicine, vol. 383, pp. 2219–2229, 2020.

[18] Athena, “Athena.ohdsi.org,” 2023. [Online]. Available: https://athena.ohdsi.org/search-terms/start. [Accessed 17 April 2023].

[19] E. Kuorttinen and M. Salmikangas, “HUS Acamedic. Virtual presentation,” 2022. [Online]. Available: https://sway.office.com/iiBUeFL0ZCF3ChY2?ref=Link. [Accessed 27 March 2023].

[20] W. McKinney, “Data Structures for Statistical Computing in Python,” Proceedings of the 9th Python in Science Conference, pp. 56–61, 2010.

[21] P. Virtanen, R. Gommers, T. E. Oliphant, M. Haberland, T. Reddy, D. Cournapeau, E. Burovski, P. Peterson, W. Weckesser, J. Bright, S. J. van der Walt, M. Brett, J. Wilson, K. J. Millman, N. Mayorov, A. R. J. Nelson, E. Jones, R. Kern, E. Larson, C. J. Carey, İ. Polat, Y. Feng, E. W. Moore, J. VanderPlas, D. Laxalde, J. Perktold, R. Cimrman, I. Henriksen, E. A. Quintero, C. R. Harris, A. M. Archibald, A. H. Ribeiro, F. Pedregosa, P. van Mulbregt and SciPy 1.0 Contributors, “SciPy 1.0: Fundamental Algorithms for Scientific Computing in Python,” Nature Methods, no. 17, pp. 261–272, 2020.

[22] M. Koskinen, J. K. Salmi, A. Loukola, M. J. Mika, J. Sinisalo, O. Carpén and R. Renkonen, “Data-driven comorbidity analysis of 100 common disorders reveals patient subgroups with differing mortality risks and laboratory correlates,” Scientific Reports, vol. 12, no. 1, pp. 1–9, 2022.

[23] D. P. Kingma and M. Welling, “Auto-Encoding Variational Bayes,” International Conference on Learning Representations, 2014.

[24] D. J. Rezende, S. Mohamed and D. Wierstra, “Stochastic Backpropagation and Approximate Inference in Deep Generative Models,” Proceedings of the 31st International Conference on Machine Learning, vol. 32, no. 2, pp. 1278–1286, 2014.

[25] F. Chollet and and others, “Keras,” 2015. [Online]. Available: https://keras.io. [Accessed 27 March 2023].

[26] L. McInnes, J. Healy and S. Astels, “hdbscan: Hierarchical density based clustering,” Journal of Open Source Software, vol. 2, no. 11, 2017.

[27] I. Goodfellow, Y. Bengio and A. Courville, Deep Learning, MIT Press, 2016.

[28] C. R. Harris, K. J. Millman, S. J. Van Der Walt, R. Gommers, P. Virtanen, D. Cournapeau, E. Wieser, J. Taylor, S. Berg, N. J. Smith, R. Kern, M. Picus, S. Hoyer, M. H. van Kerkwijk, M. Brett, A. Haldane, J. F. Del Rio, M. Wiebe, P. Peterson, P. Gérard-Marchant, K. Sheppard, T. Reddy, W. Weckesser, H. Abbasi, C. Gohlke and T. E. Oliphant, “Array programming with NumPy,” Nature, no. 585, p. 357–362, 2020.

[29] J. D. Hunter, “Matplotlib: A 2D graphics environment,” Computing in Science & Engineering, vol. 9, no. 3, pp. 90–95, 2007.

[30] M. R. Cowie, J. I. Blomster, L. H. Curtis, S. Duclaux, I. Ford, F. Fritz, S. Goldman, S. Janmohamed, J. Kreuzer, M. Leenay, A. Michel, S. Ong, J. P. Pell, M. R. Southworth, W. G. Stough, M. Thoenes, F. Zannad and A. Zalewski, “Electronic health records to facilitate clinical research,” Clinical Research in Cardiology, no. 106, pp. 1–9, 2017.

